# Unleashing the Power of Language Models in Clinical Settings: A Trailblazing Evaluation Unveiling Novel Test Design

**DOI:** 10.1101/2023.07.11.23292512

**Authors:** Qian Li, Kang Tang, Sen Li, Kaihua Zhang, Zhantao Li, Lufan Chang, Wenxue Li, Bingzhi Shen, Jia Ding, Xianjun Min

## Abstract

The realm of clinical medicine stands on the brink of a revolutionary breakthrough as large language models (LLMs) emerge as formidable allies, propelled by the prowess of deep learning and a wealth of clinical data. Yet, amidst the disquieting specter of misdiagnoses haunting the halls of medical treatment, LLMs offer a glimmer of hope, poised to reshape the landscape. However, their mettle and medical acumen, particularly in the crucible of real-world professional scenarios replete with intricate logical interconnections, remain shrouded in uncertainty. To illuminate this uncharted territory, we present an audacious quantitative evaluation method, harnessing the ingenuity of thoracic surgery questions as the litmus test for LLMs’ medical prowess. These clinical questions covering various diseases were collected, and a test format consisting of multi-choice questions and case analysis was designed based on the Chinese National Senior Health Professional Technical Qualification Examination. Five LLMs of different scales and sources were utilized to answer these questions, and evaluation and feedback were provided by professional thoracic surgeons. Among these models, GPT-4 demonstrated the highest performance with a score of 48.67 out of 100, achieving accuracies of 0.62, 0.27, and 0.63 in single-choice, multi-choice, and case-analysis questions, respectively. However, further improvement is still necessary to meet the passing threshold of the examination. Additionally, this paper analyzes the performance, advantages, disadvantages, and risks of LLMs, and proposes suggestions for improvement, providing valuable insights into the capabilities and limitations of LLMs in the specialized medical domain.

## 1 Introduction

In recent years, large language models (LLMs) have made remarkable progress in the field of natural language processing (NLP) thanks to the advancement of deep learning technology and the availability of massive text data [**Error! Reference source not found**., **Error! Reference source not found**.]. LLMs possess strong versatility and adaptability, making them applicable to a wide range of complex tasks and domains [2]. One of the significant applications is in the realm of clinical medicine, which directly impacts human life and health. Therefore, in clinical medicine, LLMs are expected to exhibit high accuracy and reliability in information processing and decision-making capabilities.

In the field of medicine, electronic health records (EHRs) serve as a crucial source of structured and unstructured data containing vital patient information. Unstructured data, such as clinical texts, primarily comprise diagnosis reports, examination results, and medical records . These clinical texts encompass a wealth of medical knowledge and patient characteristics, which play a pivotal role in medical research, clinical assistive diagnosis, and healthcare quality evaluation [3, **Error! Reference source not found**.]. However, leveraging LLMs for processing and comprehending clinical texts poses various challenges, including professional jargon, abbreviations, grammar errors, and privacy concerns. Hence, it is imperative to develop and evaluate LLMs specifically tailored for the medical domain, aiming to enhance their effectiveness and safety in medical applications.

Currently, several LLMs have been proposed specifically for the medical field. Examples include GatorTron [**Error! Reference source not found**.] and MedBERT [4], which have undergone pre-training on diverse medical datasets and subsequent evaluation on various medical NLP tasks, including clinical concept extraction, medical entity extraction, semantic text similarity, natural language reasoning, medical question answering, among others [**Error! Reference source not found**., 5]. These models have demonstrated certain advantages; however, they still exhibit certain limitations, such as small model scales, reliance on a single data source, and insufficient evaluation methodologies. Consequently, further advancements are necessary to address these limitations and enhance the performance and applicability of LLMs in the medical domain.

On the other hand, the rate of misdiagnosis during clinical treatment is a crucial issue that demands attention. Numerous studies have reported a significant variation in the misdiagnosis rates across different medical fields and diseases, ranging from 2% to 71% [6, 7]. Misdiagnosis not only poses serious harm and even fatalities to patients but also contributes to the wastage of medical resources and the occurrence of medical disputes. Therefore, the utilization of LLMs for diagnostic assistance holds the potential to reduce the misdiagnosis rate and enhance the quality and efficiency of clinical decision-making. However, it is imperative to comprehensively evaluate and verify the medical expertise of LLMs, particularly in real-world clinical scenarios. Rigorous evaluation of LLM performance within the medical profession is necessary to ensure its effectiveness and safety in medical applications.

To address these limitations, this paper proposes a novel quantitative evaluation method that assesses the medical proficiency of LLMs. Specifically, we focus on the domain of thoracic surgery and utilize diseases and clinical cases as evaluation items. To construct our evaluation framework, we gather clinically relevant questions from the Chinese National Senior Health Professional Technical Qualification Examination, encompassing various types of thoracic surgical diseases. Our assessment tool comprises multi-choice formats and case-analysis questions, uniquely designed to reflect the logical correlations of real-world clinical scenarios.

## 2 Related Works

### 2.1 The Evolution of Large Language Models

ChatGPT ^1^ as a prominent example of large language models based on generative pretrained transformer, has exhibited remarkable capabilities and extensive applicability in various domains. Its introduction in November 2022 has received significant attention and played a vital role in the advancement of large-scale language models. The Generative Pretrained Transformer (GPT) stands as an extensively trained model, leveraging vast datasets and diverse tasks, with the primary objective of establishing a versatile foundation that can be readily customized for a wide array of subsequent implementations. The inspiration behind pretraining in GPT stems from the principles of transfer learning [8, 9] in computer vision.

In more detail, early pretraining techniques, such as NNLM [10] and word2vec [11], were static and lacked adaptability to different semantic environments. To address this, dynamic pretraining techniques like BERT [12] and XLNet [13] were introduced. Besides, large language models have found a common application, such as ChatGPT, which is fine-tuned from the generative pretrained transformer and trained on a combination of text and code [**Error! Reference source not found**.]. They incorporate Reinforcement Learning from Human Feedback (RLHF) [15, 16] to align its responses with human intent, which has emerged as a promising approach for training large language models [17]. In future, the exceptional performance of LLMs has the potential to shape the training paradigms, as techniques such as reinforcement learning [15], prompt tuning [18, 19, 20], and chain-of-thought [21, 22] are employed to advance the field towards artificial general intelligence.

### 2.2 Large Language Models in Medical Domain

In clinical medicine, LLMs offer potential benefits for administrative tasks, telemedicine, clinical decision support, text summarization, efficient writing, and multilingual communication. Indeed, it is vital to address concerns regarding accuracy, timeliness, and bias in health-related information provided by LLMs. Consequently, ongoing studies are evaluating the capabilities of LLMs in different medical specialties [23-26]. For instance, Kung et al. [23] evaluated ChatGPT’s performance on the United States Medical Licensing Exam (USMLE) without specialized training or reinforcement. The results indicated a high level of concordance and insightful explanations, suggesting the potential of LLMs in medical education and clinical decision-making; Holmes et al. [24] focused on LLMs’ ability to answer radiation oncology physics questions, providing a benchmark for their performance in specialized topics relevant to the scientific and medical communities; Kasai et al. [25] evaluated ChatGPT, GPT 3, and GPT 4 ^2^ on the Japanese national medical licensing exam over a six-year period, demonstrating the potential of LLMs in the Japanese language context. Furthermore, Sarraju et al. [26] employed ChatGPT to address questions related to cardiovascular disease prevention, with responses evaluated by experienced preventive cardiology clinicians. This study highlighted the capacity of LLMs in providing appropriate medical recommendations for specific diseases, as assessed by specialists in the field.

Previous research has primarily focused on assessing the performance of large language models in general medical exams like the United States Medical Licensing Exam (USMLE) [23] and the American Society for Radiation Oncology (ASTRO) exams [24]. However, there is a necessity to explore the reasoning capabilities of LLMs in more specialized and interconnected fields within the medical domain. This paper aims to fill this gap by conducting a comprehensive evaluation of LLMs using thoracic surgery questions extracted from the Chinese National Senior Health Professional Technical Qualification Examination (Thoracic Surgery), which reflects the logical correlations of real-world clinical scenarios.

## 3 Method

### 3.1 Materials

For this research, a total of 56 questions were gathered from the Chinese National Senior Health Professional Technical Qualification Examination (Thoracic Surgery). These questions were carefully selected by experienced medical experts specializing in thoracic surgery. The set of questions comprises 30 single-choice, 20 multi-choice, and 6 case-analysis questions, aligning with the established curriculum for resident education in thoracic surgery as recommended by the National Health Commission of the People’s Republic of China. Particularly, the case analysis question type roughly simulates the complete process of a “real patient” following the routine diagnostic and treatment pathway: consultation-examination-treatment. It maximally replicates the problems encountered by doctors in real medical scenarios, including chest trauma, tumor diagnosis and treatment, as well as the coexistence of multiple diseases in the same patient. Such patients are frequently encountered in actual clinical practice, which significantly increases the difficulty of assessment and sets it apart from previous evaluation studies [23-25].

To perform a quantitative assessment of the capabilities of LLMs, we conducted a benchmark analysis utilizing the collected set of thoracic surgery questions. This analysis specifically focused on commonly used LLM APIs that were accessible as of April 28th. In order to ensure a fair comparison between the LLM models, a closed-book setting was adopted, which restricts the usage of external resources. This choice enables the results of this study to be more easily extrapolated and applied to open-book settings in future research. As for the scoring, the total score of the test is 100 points, typically consisting of 30 single-choice questions, 20 multi-choice questions, and 6 case-analysis questions. Concretely, the detailed scoring rules for each type of questions are summarized as follows,

· Single-choice questions: 1 point is awarded for a correct answer, while no points are awarded for an incorrect answer or for not selecting an option.
· Multi-choice questions: 2 points are awarded for selecting all correct options, while no points are awarded for selecting too many, too few, incorrect options, or not selecting any options.
· Case-Analysis questions: The correct answer(s) may involve one or multiple options. Each correct option selected earns 1 point, while each incorrect option selected results in a deduction of 1 point. The deduction continues until the score for that question reaches 0.

### 3.2 Models

#### Baseline Models

We evaluated five LLM APIs: ChatGPT 4^2^, ChatGPT (gpt-3.5-turbo) ^1^, Claude AI ^3^, ChatGLM-6B [27], and FastChat^4^. While the commercial LLMs don’t disclose detailed information about training data and network architectures, the open-source models provide transparency through their GitHub^5^ repositories.

All these LLMs are built on the transformer architecture [28]. Previous research has explored their capabilities on benchmark datasets like USMLE [23], ASTRO [24], JLME [25], and cardiovascular disease prevention [26].

In this study, we focus on using LLMs to tackle Thoracic Surgery questions from the Chinese National Senior Health Professional Technical Qualification Examination. By assessing their performance in this specialized domain, we aim to validate their capabilities in clinical applications.

#### Prompting and Output Formatting

Prompts are crucial in guiding LLM responses. Table 1 shows the prompts used for each question type, and Figure 2 demonstrates their concise utilization in representative examples: single-choice, multi-choice, and case-analysis questions from the Thoracic Surgery examination. This illustrates the effective use of prompts in guiding LLM responses to specific question types.

**Table 1.**
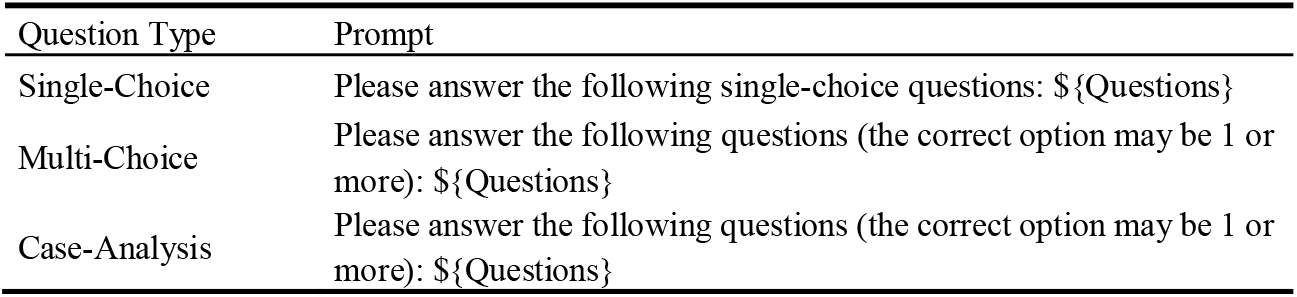
The LLM prompts used in each type of questions. ${Questions} denotes the examined question in each test.

**Fig. 2.**
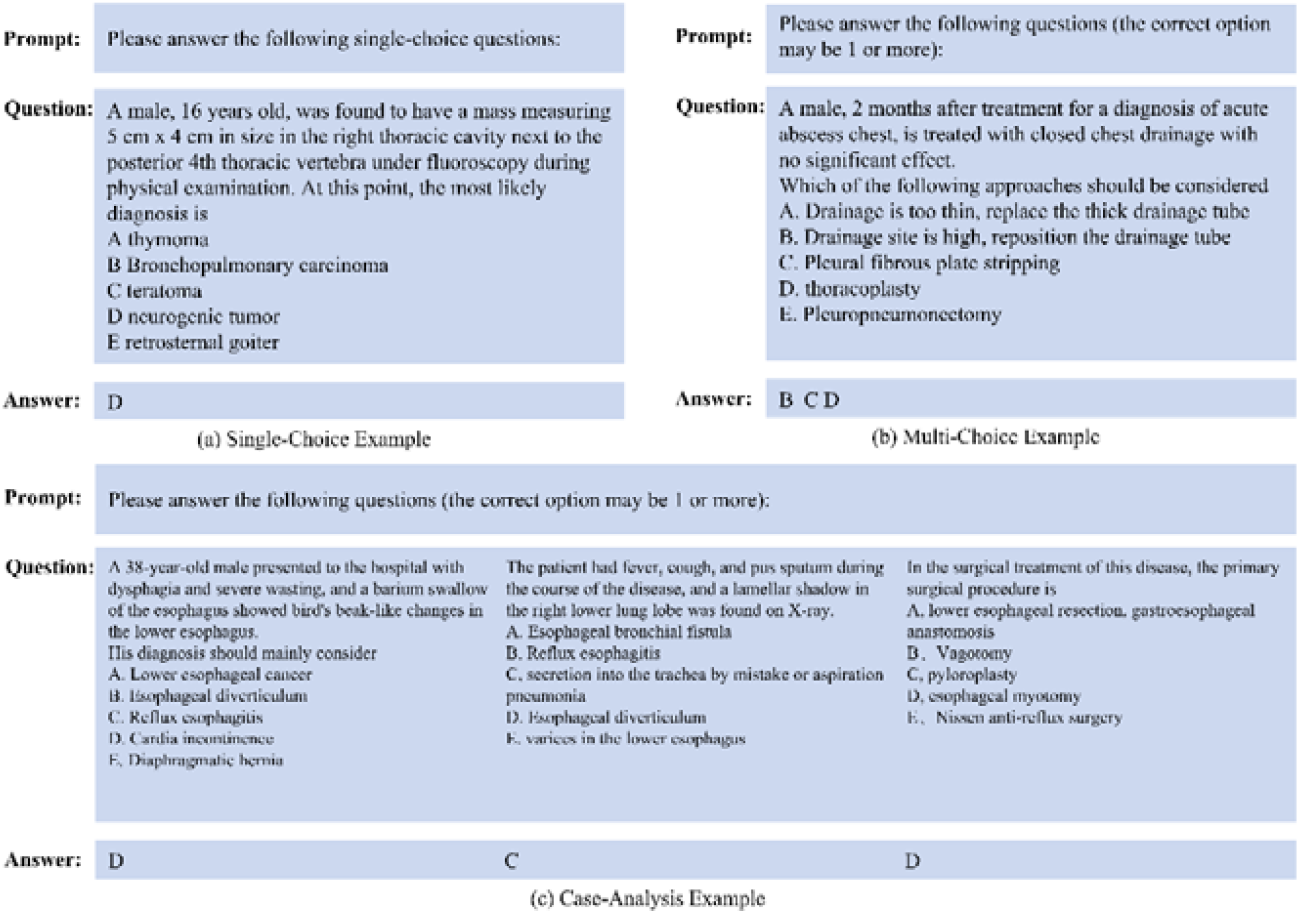
Our prompts for LLM APIs. English translations are provided here for the evaluation. Here randomly selects questions from each type of questions.

In our study, we faced the challenge of translating Chinese questions from the National Senior Health Professional Technical Qualification Examination into English. We evaluated the effectiveness of using direct prompts and example-based prompts for language model training. We conducted experiments with zero, one, and five examples. We also tested a method that instructed the model to detail its thought process step-by-step, aiming for a more structured answer. Specific example prompts can be seen in **Figure 2**.

During the evaluation phase, we utilized a parameter called “temperature” to achieve a balance between generating responses that are both creative and consistent with the context of the question. In our optimal experiments, this parameter was set to 0. However, similar to findings in previous studies on law school exams where the use of chain-of-thought prompting did not result in performance improvements [29], we did not observe any enhancements from adding intermediate steps of explanations to the LLMs in our research.

### 3.3 Evaluations

To evaluate model performance, we used exact matching in automatic evaluations, fitting for the predetermined answers of the examination. This differs from open-ended generation tasks that often require human evaluations or more advanced metrics [26]. Test scores were determined as per Section 3.1. We also compared LLMs’ scores with human scores, involving two groups: thoracic surgeons and non-experts. The surgeons had substantial thoracic surgery experience, while the non-experts held advanced degrees in technical fields but lacked thoracic surgery knowledge.

Each human participant took a two-hour closed-book test. In comparing LLMs to human scores, we calculated mean scores, evaluated score consistency, and assessed answer confidence. Consistency was measured by repeating each question three times and calculating correlations between trials.

## 4 Results

### 4.1 Overall Evaluation

Our evaluation scores (Table 2) and raw marks (Figure 3) show that the best-performing LLM model, the five-shot GPT-4, scored 48.67 out of 100. None of the models surpassed the test threshold of 60 out of 100, but GPT-3.5, GPT-4.0, Claude, and FastChat significantly outperformed random guesses, which scored just 6.6 out of 100. A considerable performance gap exists between open-source and commercial LLMs, with GPT-4 showing the highest performance among LLMs in the five-shot setting. Overall, commercial LLMs outperformed both doctor and non-expert humans, demonstrating significant advantages.

**Table 2.**
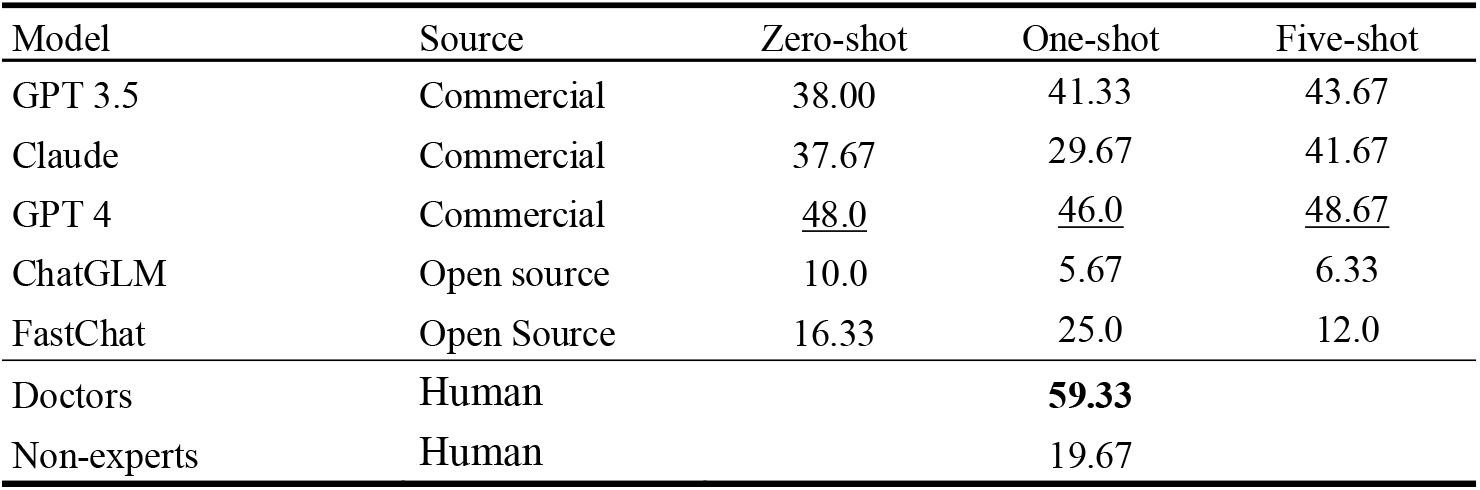
Test scores on National Senior Health Professional Technical Qualification Examination (Thoracic Surgery). The bold and underlined values are the best and second-best performance, respectively.

**Fig. 3.**
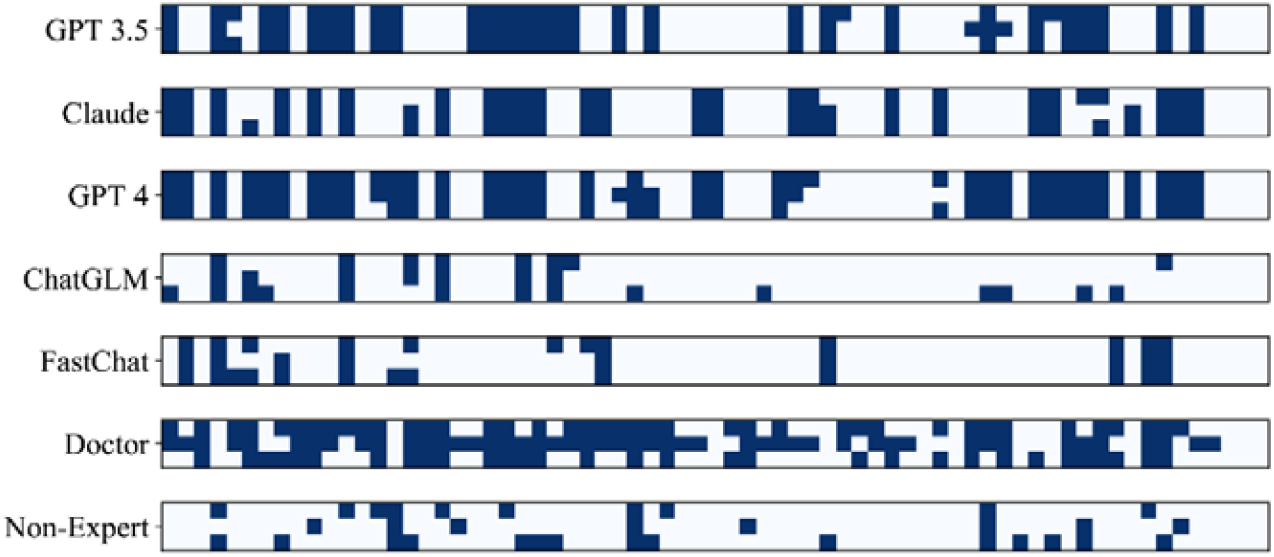
Raw marks for each test where the rows are separate tests and the columns are the test questions. Dark shaded squares represent correct answers.

We used a T-test to examine the statistical differences between model performances, reflected in p-values in Table 3. We assumed two models are equal unless the p-value is less than 0.05. Our results showed p-values greater than 0.05 for GPT 3.5, GPT 4, Claude, and Doctors, suggesting their performance is statistically equivalent and superior to other models and non-experts. Similarly, ChatGLM and FastChat, two open-source models, had p-values greater than 0.05 compared to non-experts, indicating their performance is statistically equal to non-experts. So, commercially available models perform on par with thoracic surgery experts, while open-source models align with non-experts.

**Table 3.** The p-value results of each model by T-test.

| Model | GPT 3.5 | Claude | GPT 4 | ChatGLM | FastChat | Doctors | Non-experts |
| --- | --- | --- | --- | --- | --- | --- | --- |
| GPT 3.5 | - | 0.635 | 0.266 | 2.61e-5 | 2.16e-4 | 0.055 | 4.87e-5 |
| Claude | 0.635 | - | 0.111 | 1.85e-4 | 1.21e-3 | 0.014 | 3.4e-4 |
| GPT 4 | 0.266 | 0.111 | - | 6.88e-8 | 1.17e-6 | 0.507 | 1.19e-7 |
| ChatGLM | 2.61e-5 | 1.85e-4 | 6.88e-8 | - | 0.654 | 1.32e-12 | 0.676 |
| FastChat | 2.16e-4 | 1.21e-3 | 1.17e-6 | 0.654 | - | 2.14e-10 | 0.925 |
| Doctors | 0.055 | 0.014 | 0.507 | 1.32e-12 | 2.14e-10 | - | 1.26e-12 |
| Non-experts | 4.87e-5 | 3.48e-4 | 1.19e-7 | 0.676 | 0.925 | 1.26e-12 | - |

### 4.2 Comparison between LLMs and human scores

Our study involved three thoracic surgeons and three non-expert engineers. While GPT-4 outperformed other models, it scored 10.66 less than the surgeons. The non-expert humans scored 19.67, lower than all commercial LLMs, suggesting that while commercial LLMs can logically handle thoracic surgery-related questions, a significant expertise gap compared to professional surgeons still exists.

### 4.3 Model stability

Figure 4 presents the test scores of LLMs and humans along with their corresponding error bars. When compared to doctors, LLMs exhibit lower errors, indicating that the models demonstrate greater stability than humans. Although doctors outperform LLMs in terms of performance, LLMs exhibit significantly faster response times compared to human respondents. Overall, commercial LLMs outperform humans in terms of both overall response time and accuracy.

**Fig. 4.**
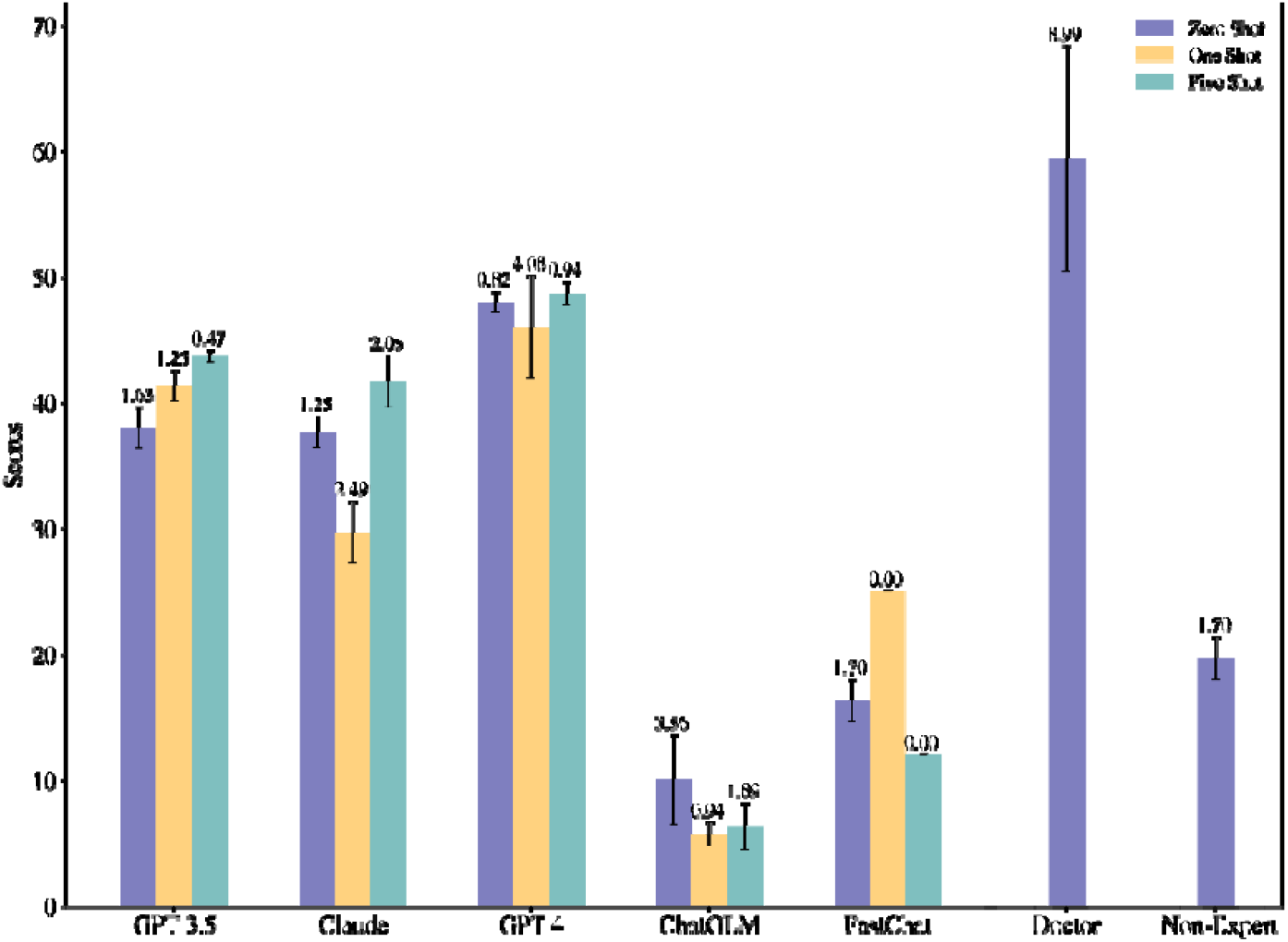
The test scores of LLMs and humans with different numbers of question examples. The error bar is indicated with their values.

### 4.4 Impact of giving examples

Our experiments used zero-shot, one-shot, and five-shot settings with prompts to evaluate LLM effectiveness. Figure 4 shows each LLM’s test scores under these settings. More examples improved some LLMs like GPT 3.5, Claude, and GPT-4, but not all. For instance, GPT 3.5 scored higher in one-shot and five-shot than in zero-shot. The best-performing LLM, GPT-4, scored lowest in one-shot but highest in five-shot, outperforming GPT-3.5’s five-shot score even in its zero-shot setting.

### 4.5 Performance on different question types

Figure 5 shows LLM proficiency across single-choice, multi-choice, and case analysis questions in a zero-shot setting. Both LLMs and humans performed better on single-choice questions and struggled with multi-choice questions. Scores were higher for case-analysis questions due to logical correlations among sub-questions. GPT-4, the top-performing LLM, matched scores on single-choice and case-analysis questions, unlike other models. Additionally, GPT-4 scored the highest in case-analysis questions among all LLMs and human experts, highlighting its deductive reasoning ability in thoracic surgery.

**Fig. 5.**
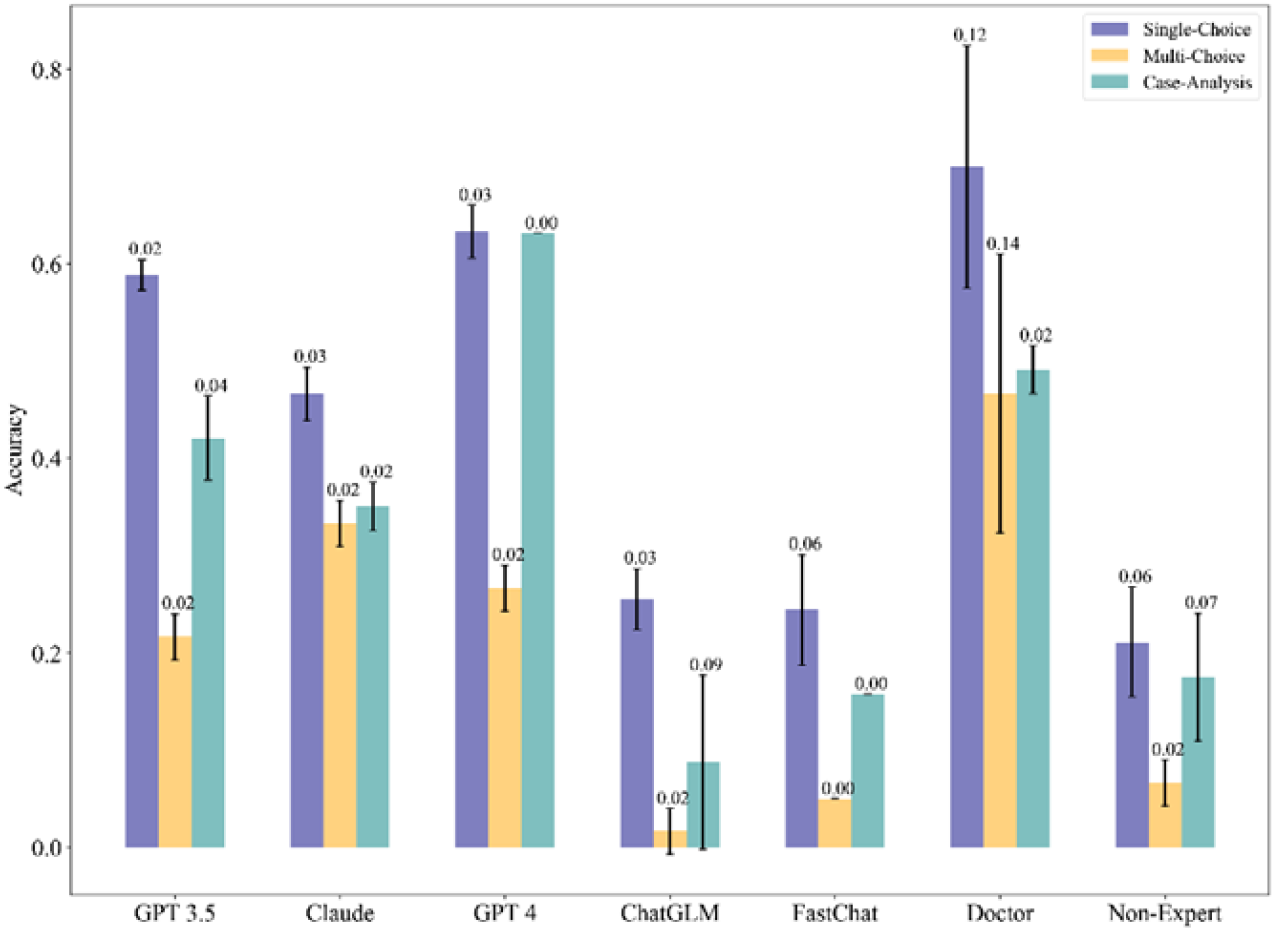
The detailed results of each type of questions. The error bar is indicated with their values.

## 5 Discussion

This study presents a novel method to quantitatively assess LLMs’ medical abilities in clinical medicine, using thoracic surgery questions as a benchmark. We curated a diverse set of clinical questions and designed a test based on the Chinese National Senior Health Professional Technical Qualification Examination. Five different LLMs were tested and evaluated by professional thoracic surgeons. Despite LLMs outperforming non-experts, with a top score of 48.67 out of 100, they failed to meet the pass threshold and lagged behind professional surgeons, primarily due to challenging question types requiring analysis, logical reasoning, and real-world clinical scenario connections. These questions required higher-order thinking skills and contextual understanding, making them more challenging for LLMs compared to previous evaluations [23, 24].

Additionally, noticeable performance differences were observed among different models. Specifically, commercial models such as GPT 3.5, GPT-4, and Claude out-performed open-source models like FastChat. This can be attributed to the higher model complexity, larger training data, and superior hardware conditions of commercial models compared to open-source models. Moreover, the commercial models also undergo real-time training and optimization through reinforcement learning and transfer learning during their application process [14]. These factors contribute to the excellent performance of commercial models on single-choice, multi-choice, and case-analysis questions, such as GPT-4.

Another factor that affects the effectiveness of the LLMs is the language of the input questions. The questions tested in this study were sourced from the Chinese National Senior Health Professional Technical Qualification Examination (Thoracic Surgery), originally written in Chinese. Since the performance of LLMs in various languages, including Chinese, lags behind their performance in English according to publicly available language proficiency rankings, it was necessary to translate the Chinese questions into English in this paper to avoid potential performance disparities due to language differences. However, it is important to note that the translation process may introduce misunderstandings and semantic losses, which could significantly constrain the performance of LLMs.

One hypothesis is that prompts may affect model performance because LLMs extract semantic information and perform model inference based on the provided prompts. But the truth is the opposite [**Error! Reference source not found**.]. Due to their focus on model stability, LLMs primarily extract key information from different prompts to generate consistent responses. Consequently, the models may not effectively capture the subtle differences between prompts or utilize the information embedded within them, which is a significant factor contributing to the inability of LLMs to pass the test. To address this issue, it is beneficial to develop specialized LLM prompt templates tailored to specific professional domains, as these domains possess more domain-specific and specialized information compared to general-purpose artificial intelligence models. Designing dedicated prompt templates can enhance the utilization of prompt information by the models, thus facilitating their inference capabilities in real-world clinical applications.

In comparing the efficiency of LLMs to that of doctors, although there are certain differences between GPT-4 and human performance, the model’s response time is significantly faster than that of humans. For instance, the response time of a local LLM, chatGLM, is about 37 minutes, whereas humans require approximately 3 hours to complete the entire examination. Therefore, considering both response capability and accuracy, once the inferential potential of LLMs in the specialized field of clinical medicine is further developed, they hold immense prospects as powerful medical assistants, addressing the issue of unequal distribution of medical resources.

This study experimentally validates the medical performance of LLMs in the field of thoracic surgery, thoroughly examining their performance across different question types and model variations, and comparing them to human test results. The results demonstrate the application potential of LLMs in the specialized medical domain. However, there are still some limitations in this study: (1) Regarding the design of the questions, due to the limited availability of publicly accessible questions from the Chinese National Senior Health Professional Technical Qualification Examination (Thoracic Surgery), this study only collected a complete set of questions and did not provide additional recombined test questions to further validate the model’s performance with different question sets; (2) In terms of answering time, the unstable nature of data transmission caused by network latency prevented us from testing the actual response time of the models. Thus, a detailed description of the LLMs’ answering time was not provided; (3) Regarding parameter validation, we set the temperature parameter in LLMs to 0, significantly unleashing the models’ inferential capabilities. However, this study did not thoroughly validate the impact of different parameter settings on model variations.

## 6 Conclusion

This paper introduces a novel method to quantitively evaluate LLMs’ abilities in specific clinical scenarios using questions from the Chinese National Senior Health Professional Technical Qualification Examination (Thoracic Surgery). Despite not passing the examination, LLMs showed potential as clinical assistants, especially with future advancements in technology, prompt engineering, model refinement, and domain-specific models. Future work includes exploring LLMs’ applications in healthcare, designing more objective testing methods, managing data specifically for LLMs in medical scenarios, and enhancing data efficiency, thus reducing costs and improving LLMs’ research and application capabilities in healthcare.

## Data Availability

All data produced in the present study are available upon reasonable request to the authors.

---

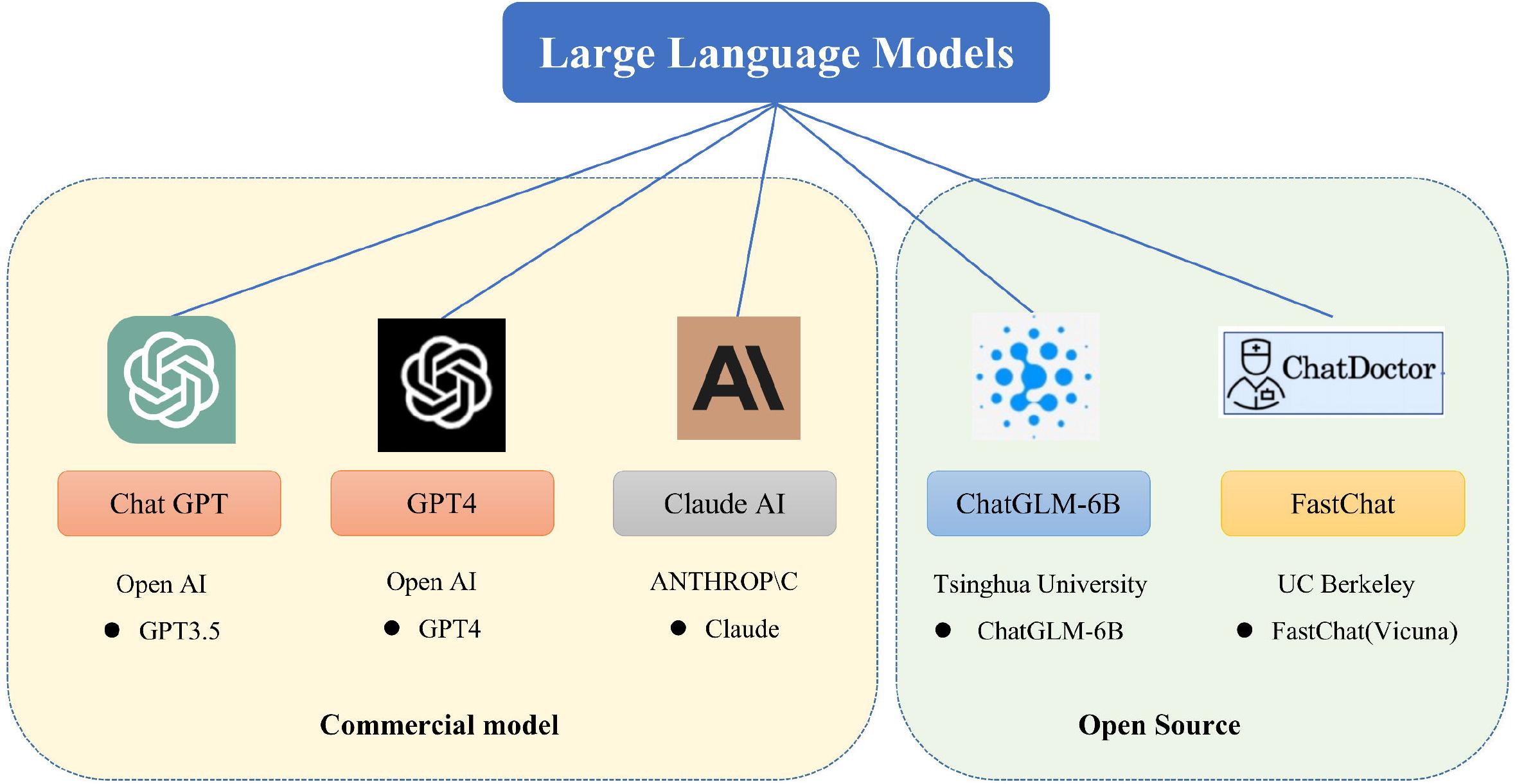

## Footnotes

1 https://openai.com/chatgpt

2 https://openai.com/gpt-4

3 http://www.anthropic.com/index/introducing-claude//www.anthropic.com/index/introducing-claude

4 https://lmsys.org/blog/2023-03-30-vicuna/

5 https://github.com/

## References

1. Wei J, Wang X, Schuurmans D, et al. Chain-of-thought prompting elicits reasoning in large language models[J]. Advances in Neural Information Processing Systems, 2022, 35: 24824–24837.

2. Shen Y, Heacock L, Elias J, et al. ChatGPT and other large language models are double-edged swords[J]. Radiology, 2023, 307(2): e230163.

3. Yang X, Chen A, PourNejatian N, et al. A large language model for electronic health records[J]. NPJ Digital Medicine, 2022, 5(1): 194.

4. Liu N, Hu Q, Xu H, et al. Med-BERT: A pretraining framework for medical records named entity recognition[J]. IEEE Transactions on Industrial Informatics, 2021, 18(8): 5600–5608.

5. Anmol Arora and Ananya Arora. The promise of large language models in healthcare. The Lancet, 401(10377):641, 2023.

6. Jonathan A Edlow and Peter J Pronovost. Misdiagnosis in the emergency department: time for a system solution. JAMA, 329(8):631–632, 2023.

7. Ohad Oren, Ron Blankstein, and Deepak L Bhatt. Addressing imaging pitfalls to reduce cardiovascular disease misdiagnosis in patients with breast cancer following reconstruction. JAMA cardiology, 7(2):123–125, 2022.

8. Xiang Yu, Jian Wang, Qing-Qi Hong, Raja Teku, Shui-Hua Wang, and Yu-Dong Zhang. Transfer learning for medical images analyses: A survey. Neurocomputing, 489:230–254, 2022.

9. Fuzhen Zhuang, Zhiyuan Qi, Keyu Duan, Dongbo Xi, Yongchun Zhu, Hengshu Zhu, Hui Xiong, and Qing He. A comprehensive survey on transfer learning. Proceedings of the IEEE, 109(1):43–76, 2020.

10. Yoshua Bengio, Réjean Ducharme, and Pascal Vincent. A neural probabilistic language model. Advances in neural information processing systems, 13, 2000.

11. Tomas Mikolov, Kai Chen, Gregory S. Corrado, and Jeffrey Dean. Efficient estimation of word representations in vector space. In International Conference on Learning Representations, 2013.

12. Jacob Devlin, Ming-Wei Chang, Kenton Lee, and Kristina Toutanova. BERT: Pre-training of deep bidirectional transformers for language understanding. In Proceedings of the 2019 Conference of the North American Chapter of the Association for Computational Linguistics: Human Language Technologies, Volume 1(Long and Short Papers), pages 4171–4186, Minneapolis, Minnesota, June 2019. Association for Computational Linguistics.

13. Zhilin Yang, Zihang Dai, Yiming Yang, Jaime Carbonell, Ruslan Salakhutdinov, and Quoc V. Le. XLNet: Generalized Autoregressive Pretraining for Language Understanding. Curran Associates Inc., Red Hook, NY, USA, 2019.

14. Katharine Sanderson. Gpt-4 is here: what scientists think. Nature, 615(7954):773, 2023.

15. Paul F Christiano, Jan Leike, Tom Brown, Miljan Martic, Shane Legg, and Dario Amodei. Deep reinforcement learning from human preferences. Advances in neural information processing systems, 30, 2017.

16. Nisan Stiennon, Long Ouyang, Jeffrey Wu, Daniel Ziegler, Ryan Lowe, Chelsea Voss, Alec Radford, Dario Amodei, and Paul F Christiano. Learning to summarize with human feedback. Advances in Neural Information Processing Systems, 33:3008–3021, 2020.

17. Long Ouyang, Jeffrey Wu, Xu Jiang, Diogo Almeida, Carroll Wainwright, Pamela Mishkin, Chong Zhang, Sandhini Agarwal, Katarina Slama, Alex Ray, et al. Training language models to follow instructions with human feedback. Advances in Neural Information Processing Systems, 35:27730–27744, 2022.

18. Xiao Liu, Kaixuan Ji, Yicheng Fu, Weng Tam, Zhengxiao Du, Zhilin Yang, and Jie Tang. P-tuning: Prompt tuning can be comparable to fine-tuning across scales and tasks. In Proceedings of the 60th Annual Meeting of the Association for Computational Linguistics (Volume 2: Short Papers), pages 61–68, 2022.

19. Pengfei Liu, Weizhe Yuan, Jinlan Fu, Zhengbao Jiang, Hiroaki Hayashi, and Graham Neubig. Pre-train, prompt, and predict: A systematic survey of prompting methods in natural language processing. ACM Computing Surveys, 55(9):1–35, 2023.

20. Kaiyang Zhou, Jingkang Yang, Chen Change Loy, and Ziwei Liu. Learning to prompt for vision-language models. International Journal of Computer Vision, 130(9):2337–2348, 2022.

21. Zhuosheng Zhang, Aston Zhang, Mu Li, and Alex Smola. Automatic chain of thought prompting in large language models. In International Conference on Learning Representations, 2023.

22. Jason Wei, Xuezhi Wang, Dale Schuurmans, Maarten Bosma, Ed Chi, Quoc Le, and Denny Zhou. Chain of thought prompting elicits reasoning in large language models. In Advances in Neural Information Processing Systems, 2022.

23. Tiffany H Kung, Morgan Cheatham, Arielle Medenilla, Czarina Sillos, Lorie De Leon, Camille Elepaño, Maria Madriaga, Rimel Aggabao, Giezel Diaz-Candido, James Maningo, et al. Performance of chatgpt on usmle: Potential for ai-assisted medical education using large language models. PLoS digital health, 2(2):e0000198, 2023.

24. Jason Holmes, Zhengliang Liu, Lian Zhang, Yuzhen Ding, Terence T Sio, Lisa A McGee, Jonathan B Ashman, Xiang Li, Tianming Liu, Jiajian Shen, et al. Evaluating large language models on a highly-specialized topic, radiation oncology physics. arXiv preprint arXiv:2304.01938, 2023.

25. Jungo Kasai, Yuhei Kasai, Keisuke Sakaguchi, Yutaro Yamada, and Dragomir Radev. Evaluating gpt-4 and chatgpt on japanese medical licensing examinations. arXiv preprint arXiv:2303.18027, 2023.

26. Ashish Sarraju, Dennis Bruemmer, Erik Van Iterson, Leslie Cho, Fatima Rodriguez, and Luke Laffin. Appropriateness of cardiovascular disease prevention recommendations obtained from a popular online chat-based artificial intelligence model. JAMA, 329(10):842–844, 2023.

27. Zhengxiao Du, Yujie Qian, Xiao Liu, Ming Ding, Jiezhong Qiu, Zhilin Yang, and Jie Tang. Glm: General language model pretraining with autoregressive blank infilling. In Proceedings of the 60th Annual Meeting of the Association for Computational Linguistics (Volume 1: Long Papers), pages 320–335, 2022.

28. Kai Han, An Xiao, Enhua Wu, Jianyuan Guo, Chunjing Xu, and Yunhe Wang. Transformer in transformer. Advances in Neural Information Processing Systems, 34:15908–15919, 2021.

29. Jonathan H Choi, Kristin E Hickman, Amy Monahan, and Daniel Schwarcz. Chatgpt goes to law school. Available at SSRN, 2023.

30. Shrivastava D, Larochelle H, Tarlow D. Repository-level prompt generation for large language models of code[C]//International Conference on Machine Learning. PMLR, 2023: 31693–31715.

